# Predictive ALS survival using ALSFRS-R slope & NfL: insights from the ALS/MND Natural History Consortium data and biofluid collection

**DOI:** 10.64898/2026.08.06.26359910

**Authors:** Andres Arguedas, Danni Li, Kelly Duffy, Annette Xenopoulos-Oddsson, James Wymer, Terry Heiman-Patterson, Ghazala Hayat, Mehdi Ghasemi, Tawfiq Al-Lahham, Senda Ajroud-Driss, Nicholas Olney, Ximena Arcila-Londono, Kelly Gwathmey, Alex Sherman, Mark Fiecas, Erjia Cui, David Walk

## Abstract

**Background:** Amyotrophic lateral sclerosis (ALS) is a rare neurodegenerative disease with no known cure. Disease progression in people living with ALS is heterogeneous, hindering personalized treatment development. The current gold standard for measuring disease progression in ALS, the ALS Functional Rating Scale - Revised (ALSFRS-R), is widely used but based on subjective measurements. Blood-based neurofilament light (NfL) has been studied as a diagnostic and prognostic biomarker but less information exists on its utility as a disease progression biomarker.

**Methods:** We present results from blood draws of 300 participants in the FDA-funded Clinic-Based Multi-Site ALS Natural History and Biofluid study of the ALS Natural History Consortium (NHC). Plasma NfL levels were measured and analyzed against different disease progression metrics based on the ALSFRS-R.

**Results:** NfL levels were found to be correlated with the ALSFRS-R average rate of change (r=−0.53, 95% CI −0.62 to −0.42). This association differed at a cutoff value of 61 pg/mL, with stronger correlations below this cutoff (r=−0.51 vs r=−0.18). Survival differed stratifying by this cutoff value, with participants under the cutoff having higher survival probabilities. The predictive value of NfL when predicting time to death was higher compared with the first ALSFRS-R across different event horizons. A model including both was better when predicting events up to 2 years after diagnosis.

**Conclusions:** These results highlight the utility of NfL as a disease progression biomarker in ALS alongside ALSFRS-R based disease progression metrics. The cutoff value can aid in clinical trial stratification, pragmatic trial planning, and clinical care.

**KEY MESSAGES:** *What is already known on this topic:* Blood-based neurofilament light (NfL) has previously been identified as a diagnostic and prognostic biomarker in ALS, but its use as a disease progression biomarker has been less studied.

*What this study adds:* This manuscript presents findings related to the utility of NfL as a disease progression biomarker in ALS from the current largest natural history cohort of people living with ALS. Specifically, it shows the added utility of baseline NfL values in predicting both disease progression and survival.

*How this study might affect research, practice, or policy:* Evidence in favor of the use of NfL as a disease progression biomarker can impact the design of clinical trials, helping with patient stratification, and allowing for better tailored treatments for people living with ALS. NfL is also being collected more frequently in standard clinical care, making a cutoff value for disease progression useful for disease management and patient planning.

## INTRODUCTION

Amyotrophic lateral sclerosis (ALS) is a progressive neurological disorder which currently does not have a highly effective treatment for most patients [1,2]. The Amyotrophic Lateral Sclerosis Functional Rating Scale - Revised (ALSFRS-R) [3] is routinely measured during clinical visits and is widely used in the field of ALS as a measure of disease progression and function. The change in ALSFRS-R over time is commonly used as the primary endpoint in clinical trials in ALS. There is a recognized need for biomarkers of disease progression to supplement this functional outcome measure in ALS, particularly as a secondary outcome for clinical trials [4–6]. Neurofilament light (NfL) is an axonal protein in large myelinated nerve fibers. Elevated NfL in blood is a biomarker of neurodegeneration and is present in many neurological diseases, including ALS [7,8]. Many studies have established the utility of blood-based NfL as a prognostic biomarker in ALS, and one drug has been approved for use in ALS in part based upon evidence that it reduced NfL levels in cerebrospinal fluid (CSF) and plasma [9–12]. In part because of this, blood biomarkers have garnered wider attention from the ALS community [13].However, as of yet there is little evidence of NfL’s utility as a disease progression biomarker in the natural history of ALS.

Other blood-based biomarkers have emerged as potentially useful as prognostic, progression, or phenotypic biomarkers in ALS. Cardiac Troponin T (cTnT) is expressed in diseased and regenerating skeletal muscle [14]. A limited number of case reports and a case–control study suggest that cTnT is elevated in ALS in the absence of evidence of cardiac damage [15,16]. A recent study showed that serum cTnT levels increase over time and correlate with disease severity as measured with ALSFRS-R [17]. cTnT may serve as a proxy of lower motor neuron involvement. In addition, a recent study showed that plasma p-tau 181, which has hitherto been considered a biomarker for Alzheimer’s disease, may be a novel biomarker of LMN dysfunction in ALS [18].

The Clinic-Based Multi-Site ALS Natural History and Biofluid study collects longitudinal clinical data and biofluids from several member sites of the ALS Natural History Consortium (ALS NHC) in the United States. Participants agree to sharing de-identified clinical datasets [19] and have the option to provide blood collections. In selected individuals, blood is collected longitudinally for up to 12 months.

In this report, we present the correlations between baseline blood-based NfL, cTnT, and p-Tau181 obtained at enrollment in the ALS NHC with clinical phenotype, ALSFRS-R progression, and survival.

## MATERIALS AND METHODS

This study uses data collected from the Clinic-Based Multi-Site ALS Natural History and Biofluid study, which collects longitudinal clinical data and blood from member sites of the ALS Natural History Consortium (NHC) in the United States. Starting in 2024 blood was collected with consent at enrollment from 300 people living with ALS in the NHC dataset and analyzed for neurofilament light (NfL), plasma phosphorylated tau-^181^ (pTau^181^), and cardiac troponin T (cTnT). Given the nature of the data both prevalent (enrolled more than 90 days after diagnosis) and incident cases (enrolled within 90 days after diagnosis) were included in this cohort. The median time of biomarker collection was 196 days after diagnosis (IQR = (60, 526)).

EDTA whole blood was collected and transported refrigerated overnight to the central lab at the University of Minnesota Advanced Research and Diagnostic Lab (ARDL). Upon arrival, the blood samples were centrifuged at room temperature to produce plasma samples, which were then aliquoted and stored at −80 celsius for long-term storage. EDTA plasma was used to measure NfL and p-tau 181 using commercially available SiMoA assays on an HD-X analyzer (Quanterix, MA) in the ARDL; serum samples were used to measure cTnT using the Roche high-sensitivity Troponin Gen5 assay. Because we ran plasma NfL and p-tau 181 samples in two batches with different reagent lots, we harmonized the results by including the same set of 20 samples in both batches. cTnT was measured using the Roche Cobas instrument, which is a random-access instrument and uses reagents without batch effects; therefore, harmonization was not necessary for the cTnT results.

The average rate of change of the ALSFRS-R scores was computed based on a mixed linear model with normal errors. The total ALSFRS-R score was the response variable, and the model included a fixed effect for time and a random intercept and slope. For every participant all ALSFRS-R scores collected over time were included in this model. Participants with no observed ALSFRS-R scores were thus excluded from the analysis. The predicted random slope from this model for each participant was used as the average rate of change of the ALSFRS-R score. The average rate of change of the ALSFRS-R was compared to biomarker values using Spearman’s correlation, with a significance test to assess the potential association between these variables. A multiple linear regression model was fitted using the average rate of change of the ALSFRS-R as the outcome variable, and the three biomarkers as covariates, to determine the association of these biomarkers jointly with disease progression.

To determine a cut-off value of NfL for analyzing disease progression, a changepoint model was fit. A linear spline model [20] with the estimated ALSFRS-R slope as a response variable was fit, using both the NfL values as well as the maximum between 0 and the difference between the NfL value and the cut-off value as predictors. This approach allows for a continuous line to be fit with a change of slopes at the changepoint. To determine the cut-off value, a grid of 1,000 values uniformly distributed across the observed NfL values was chosen. A model was fit at each of these cut-off values, and the one that produced the model with the smallest Akaike’s Information Criterion (AIC) [21] value was chosen as the cut-off.

The relationship between biomarker values and time-to-death were assessed using Kaplan-Meier curves and Cox proportional hazards (PH) models. For the Kaplan-Meier curve, a stratified analysis was conducted, separating participants based on the median NfL value of the analytic sample. These groups were compared for differences by fitting a Cox PH model including an indicator for if NfL values were above or below the cut-off. Additional Cox models were fitted using the first ALSFRS-R score and its slope to assess the added value of biomarkers compared to usual disease progression measurements. Due to the measurement scale of the biomarkers, coefficients and hazard rates are reported for 10 unit increases.

The predictive performance of the Cox models was assessed through the dynamic AUC [22,23], and C-index [24]. At a given time point of interest and with two randomly selected participants, one with an event and the other without, the dynamic AUC is calculated as the probability that the model will correctly assign the participant with the event a higher risk than the one without an event. These values can be used to compare the predictive capabilities of different models at various time points of interest. In this study we calculated AUC values for each model every six months after diagnosis. Similarly, the C-Index quantifies the probability that, out of two randomly chosen participants, the one with a higher estimated risk will have a shorter survival time than the one with a lower risk. For both metrics a value of 1 indicates perfect prediction, while a value of 0 indicates completely imperfect prediction, while 0.5 indicates a result equivalent to a coin toss. Higher values indicate better predictive performance of a model.

The data and biosample collection for this study are approved by the University of Minnesota IRB as the IRB of record. The current IRB number is sIRB 00019204. Patients seen at ALS multidisciplinary clinics at participating centers are offered participation, consisting of sharing of de-identified clinical data and an optional blood collection. The data used in the analysis of this manuscript is available upon request for valid research purposes from the senior author.

## RESULTS

At the time of manuscript preparation, 556 blood collections had been completed at the time of study enrollment, of whom 300 have had plasma biomarker assays to date. Biomarker and initial phenotype data were available for 296 participants. ALSFRS-R slope and survival data were available from 214 of these. The CONSORT diagram is provided in Figure 1. Table 1 presents characteristics at baseline for participants in the analytic sample.

**Table 1.** Sociodemographic and disease characteristics for participants at study baseline. Summary statistics are Median (IQR) for continuous variables and n (%) for categorical variables.

| Variable | Summary Measure |
| --- | --- |
| Age at diagnosis (years) | 64 (58, 71) |
| Time from symptom onset to enrollment (months) | 16 (11, 35) |
| Time from diagnosis to enrollment (months) | 3 (1, 7) |
| Time from diagnosis to biomarker collection (months) | 8 (2, 21) |
| Time from diagnosis to first ALSFRS-R (months) | 1 (0, 4) |
| Female | 102 (48%) |
| Non-Hispanic/Latino | 205 (99%) |
| White | 190 (90%) |
| Limb onset | 156 (75%) |
| Number of visits | 6 (4, 11) |

**Figure 1.**
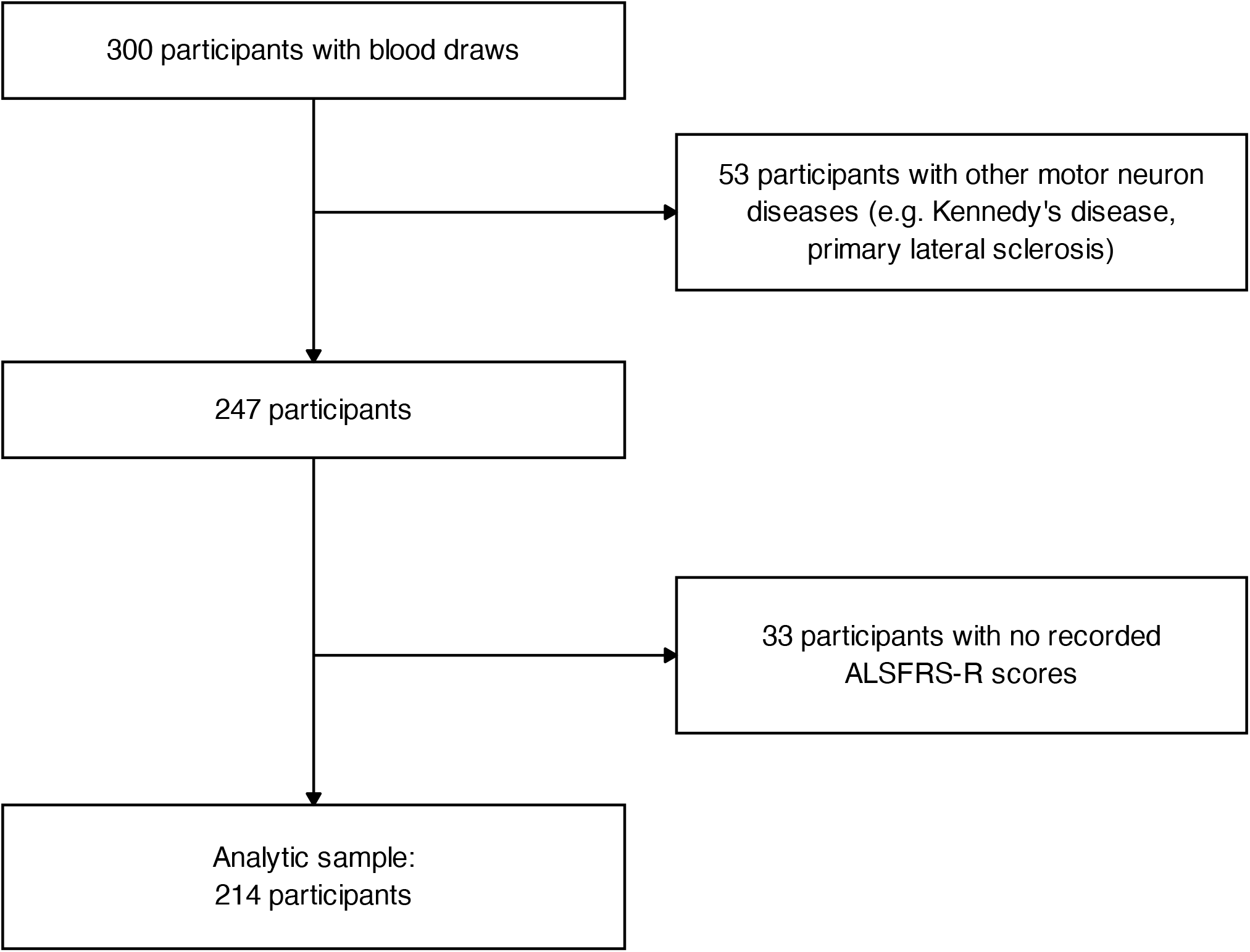
Consort diagram of inclusion and exclusion criterion for the analytic sample.

Figure 2 shows a scatterplot of the initial NfL values and the estimated ALSFRS-R slope, which measures the monthly average change in the ALSFRS-R. There is a moderate negative correlation between the NfL values and the ALSFRS-R slope (r=−0.53, 95% CI −0.62 to −0.42), indicating that, on average, participants with higher baseline NfL values have more negative ALSFRS-R slopes. A cut-off value for NfL of 61 pg/mL was estimated based on a changepoint model with the lowest AIC at that value. The association between NfL levels and ALSFRS-R average rate of change differed based on this cutoff value. The correlation between these two measures was higher below the cutoff value (r= −0.54) compared to above the cutoff (r= −0.18) The average ALSFRS-R rate of change was also comparatively lower for participants under the cut-off compared to those above it (−0.51 and −1.10 respectively, 95% CI for the difference 0.45 to 0.72).

**Figure 2.**
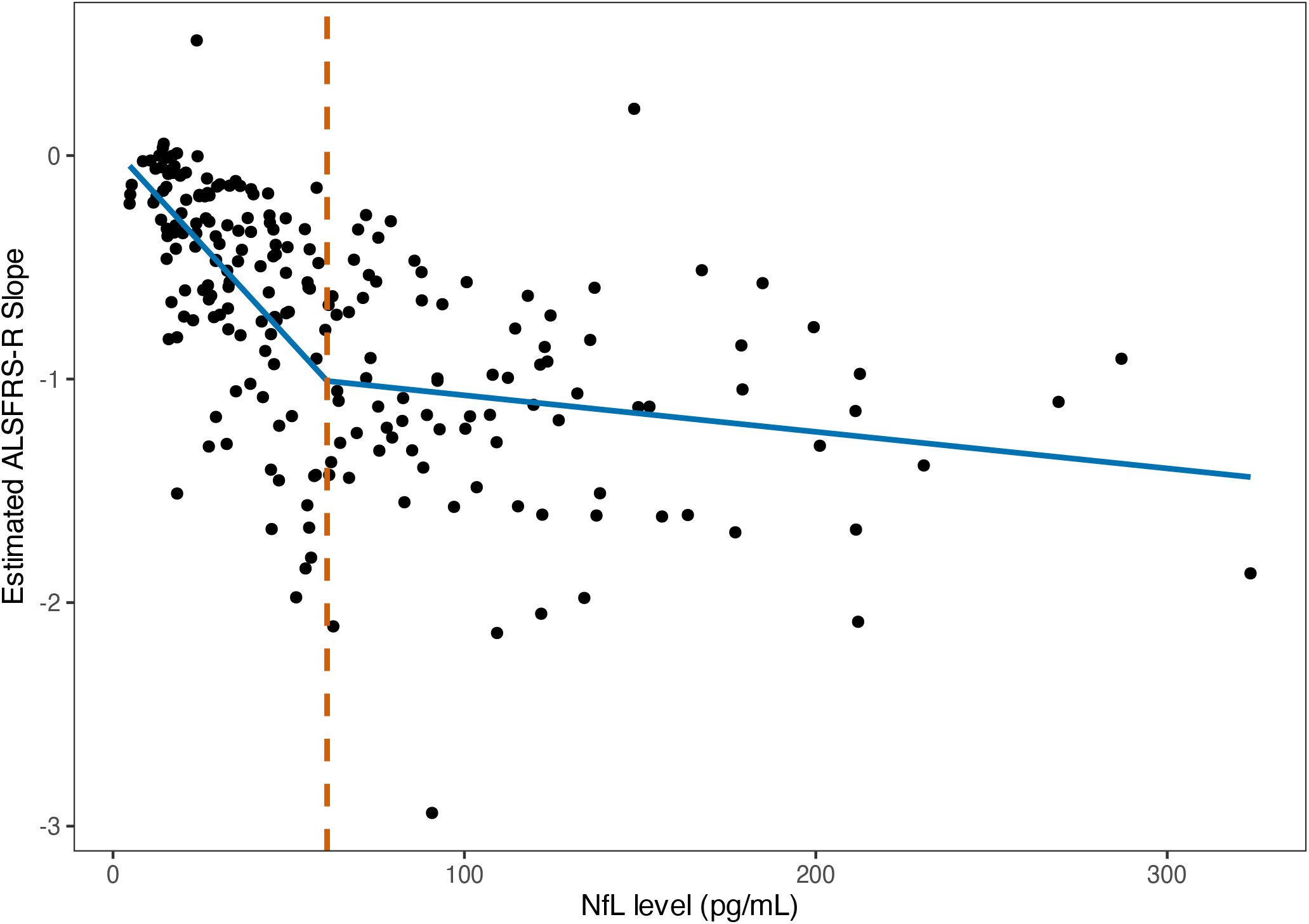
Association of NfL values with the estimated ALSFRS-R slope. The orange dashed line corresponds to the estimated cut-off value of 61 pg/ml. The blue line corresponds to the estimated regression line with a changepoint at the cut-off value.

Neither pTau181 (r=−0.11, 95% CI −0.14 to 0.12) nor cTnT (r=0.06, 95% CI −0.08 to 0.21) showed a notable correlation with ALSFRS-R slope. When fitting a multiple linear regression model using the ALSFRS-R slope as the outcome and all three biomarkers as predictors, NfL shows a negative effect for every 10 pg/mL increase (β=−0.05, 95% CI −0.06 to −0.04), while pTau181 and cTnT show no effect (β=−0.01, 95% −0.03 to 0.02; and β=0.01, 95% CI −0.02 to 0.04, respectively).

Figure 3 presents the Kaplan-Meier (KM) curve for all participants in the analytic sample. This KM curve reaches the median survival probability about 4 years after diagnosis. A stratified analysis based on the cutoff NfL value (61 pg/mL) is also presented in Figure 3, with a KM curve for participants with NfL values at or above the cut-off value, and another curve for those with values below the cut-off value. These two curves do not intersect and are clearly differentiated both from each other and from the overall KM curve, with the curve for those with NfL values below the cut-off still not reaching the median survival probability at 6 years after diagnosis, while those with NfL values at or above the cut-off value reach a survival probability of 0.5 less than 2 years after diagnosis. When fitting a Cox PH model using an indicator for NfL values above or below the cut-off, participants with NfL values over the cut-off had considerably higher hazards (HR=5.21, 95% 3.09 to 8.79). When fitting a Cox PH model with all three biomarkers, a change of 10 pg/mL in NfL values increased the overall risk of death by 20% (HR=1.21, 95% CI 1.16 to 1.25). These associations were not observed for pTau181 (HR=1.03, 95% CI 0.96 to 1.11) or cTnT (HR=1.03, 95% CI 0.96 to 1.11).

**Figure 3.**
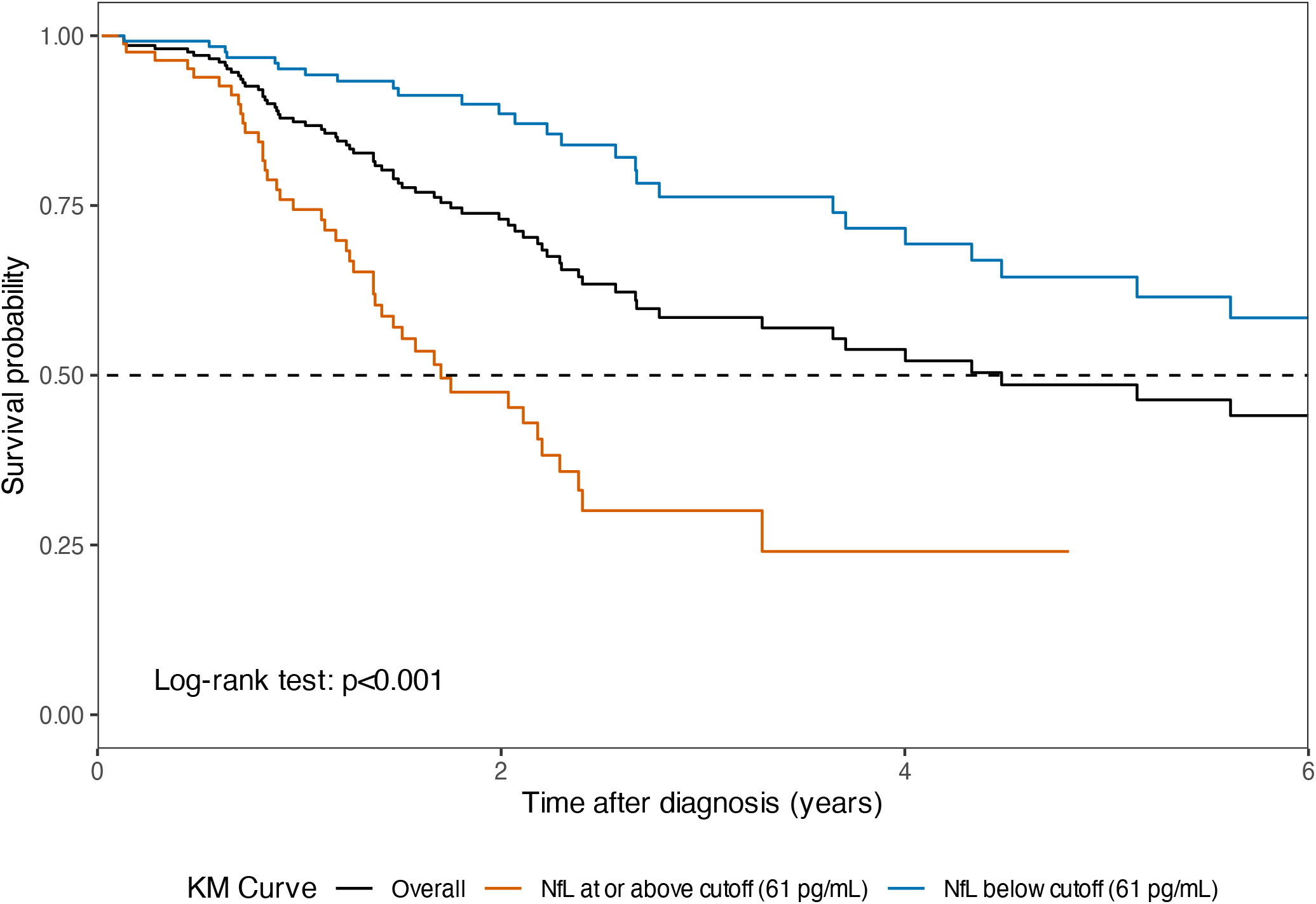
Kaplan Meier curve of overall ALS survival within the analytic sample. The black line presents the curve for the complete analytic sample, while the red and blue lines denote the KM curves stratified by NfL values above or below the estimated cut-off value of 61 pg/ml, respectively.

Figure 4 presents the dynamic AUCs for mortality comparing models with baseline NfL values and the first observed ALSFRS-R score. The model with only the baseline NfL value outperforms the model with only the first observed ALSFRS-R score, indicating better predictive capability.

**Figure 4.**
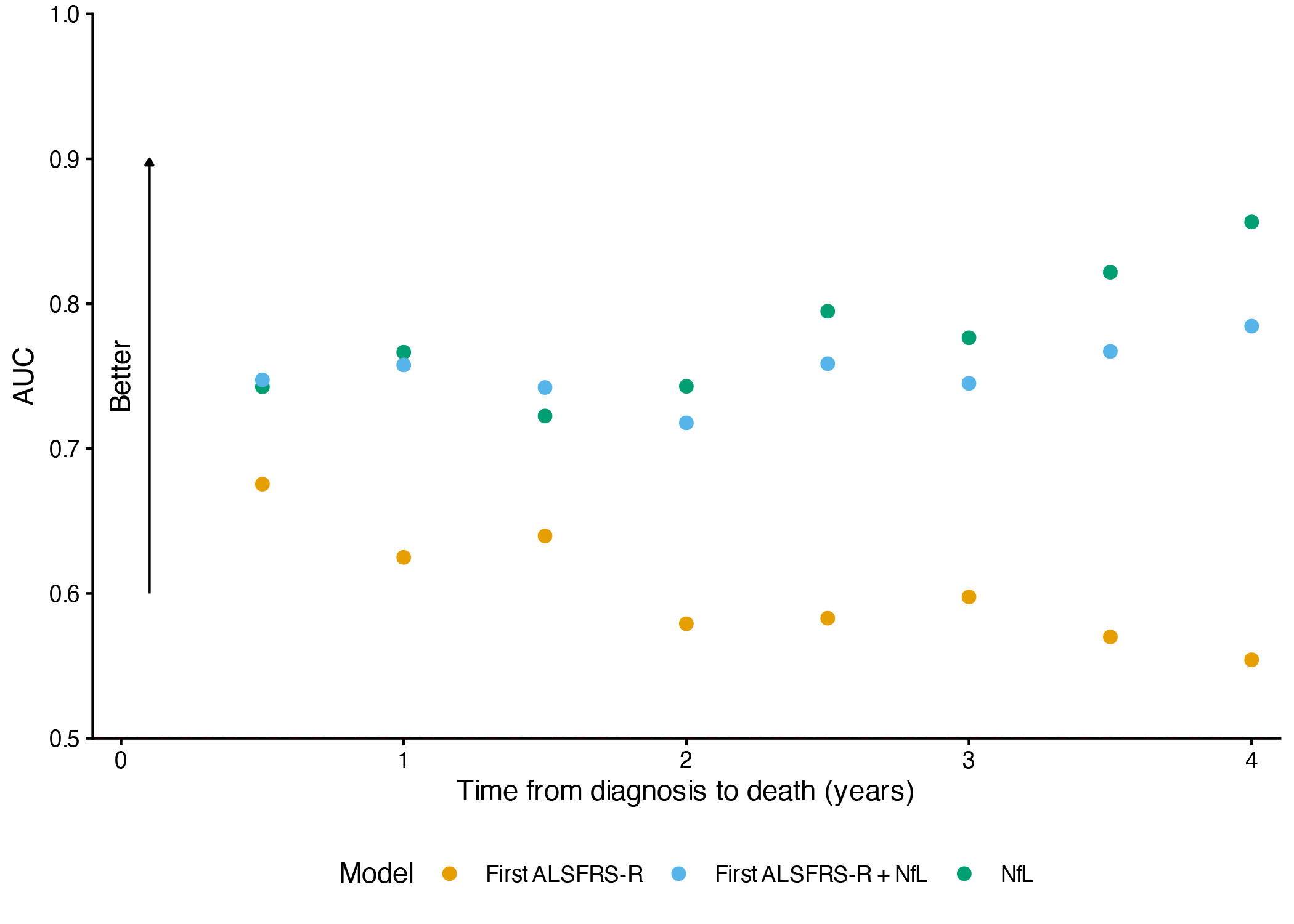
Dynamic AUC for survival prediction models using baseline NfL values and ALSFRS-R related variables every 6 months after diagnosis (time 0). Models including the baseline NfL value and the first observed ALSFRS-R total score are presented.

When using both variables in one model, the combined model outperforms the models with each of the variables separately before 2 years, with the model using only NfL alone outperforming it after 2 years. An additional analysis utilizing the estimated rate of change of the ALSFRS-R scores was also conducted (Figure S1). Models including the average rate of change of the ALSFRS-R score, either by itself or with the baseline NfL value, outperform a model with only NfL after 6 months. Including NfL shows an improvement in the predictive capabilities of the model when compared to a model with only the average rate of change of the ALSFRS-R score before 2 years.

When measuring overall predictive capabilities for survival using the C-index, the model with the baseline NfL value (C-index: 0.87) performs better than the one with the first recorded ALSFRS-R score (C-index: 0.80). The model’s predictive capabilities increased when including both the NfL value and the first recorded ALSFRS-R score (C-index: 0.89). A model with the ALSFRS-R slope (C-index: 0.75) also shows predictive improvement when adding NfL values (C-index: 0.87).

## DISCUSSION

We show that a single NfL value correlates well with ALSFRS-R slope (especially when NfL is less than 61 pg/mL) and is predictive of survival. The high AUC values across different time points and an overall high C-index strengthen the utility of a single baseline NfL value for predicting survival throughout a person’s disease course. Throughout all time points a model with only NfL outperforms a model with only the first observed ALSFRS-R score. A model using both variables outperforms models with variables used individually when predicting death within 2 years of diagnosis. Including the average rate of change of the ALSFRS-R score yields overall better predictive performance than using the baseline NfL value by itself. A model using both variables outperformed the model using only the average rate of change of the ALSFRS-R score until 1.5 years after diagnosis, after which using the model with only the average rate of change of the ALSFRS-R score is best. This indicates that NfL may help with predictions soon after diagnosis, even in the hypothetical case when the full longitudinal disease progression trajectory is known. pTau181 and cTnT did not contribute to survival prediction or correlate with ALSFRS-R slope as a measure of disease progression rate.

The association between NfL and prognosis is recognized, and there have been some efforts to use NfL to stratify both diagnosis and prognosis. Cut-off values have been constructed based on survival outcomes [25,26], progression via the ALSFRS-R slope [11], or directly from NfL values [27]. For the cut-offs based on survival, these have been reported around 80-95 pg/mL, while the cut-offs based on disease progression were obtained for tertiles and corresponded to 65, 114, and 160, approximately. One approach created 10 groups based on Z-scores, with a median of 112 pg/mL.

In our cohort, the relationship between ALSFRS-R decline and NfL is non-linear; specifically, there appears to be a change in the rate of this relationship at a plasma NfL level of about 61 pg/ml. Insofar as higher levels are associated with a similarly rapid decline in functional scores, we used that cutoff to stratify by survival and found a clear difference. Previous proposed cut-off values either segmented disease progression and then defined cut-off values or classified based only on NfL values without explicitly including disease progression measures. Our proposed cut-off is lower than the aforementioned values and was constructed based on both disease progression and NfL values directly. This aids in segmenting patients, since those above the cut-off value had faster disease progression on average compared to those below the cut-off while having a less pronounced change as NfL values increased. This cut-off value might be used to stratify patients in clinical trials to better address heterogeneous disease progression. Nevertheless, since there is a lack of standardization when measuring blood-based NfL levels [28], an equivalent value for this cut-off should be used for different samples.

Survival prediction in ALS is useful for personal prediction, designing clinical trial inclusion criteria, and assessment of real-world data. Our data demonstrate that a single NfL value provides additional survival predictive value when ALSFRS-R data are available. Over time, for those who survive 2 years or more, ALSFRS-R slope is sufficient to predict further survival. It is important to highlight that the use of the full longitudinal data for the ALSFRS-R from participants to compute the average rate of change of the ALSFRS-R score requires using data in the future to predict at baseline. This implies that the results from our predictions using the average rate of change of the ALSFRS-R are optimistic. More appropriate models, such as those using a landmark approach, should be used in practice for properly obtaining predictions. It is also important to note that our survival prediction includes NfL values obtained from both recently diagnosed as well as prevalent cases, which introduces a bias. Insofar as NfL has been collected from our cohort since early 2024 and has only been in routine clinical use for a similar duration, this is a necessary limitation of such work at this time.

Subsequent work will include longer clinical follow up of this cohort and additional patient level characteristics in the model, enabling more complex models to be fit to the data. The addition of longitudinal biomarker data will also enhance our understanding of NfL, cTnT, and p-tau181 as biomarkers of disease progression.

## Data Availability

The data used in the analysis of this manuscript is available upon request for valid research purposes from the senior author.

## ACKNOWLEDGMENTS

This project was made possible by grant number R01FD007630 from FDA’s Office of Orphan Products Development. Its contents are solely the responsibility of the authors and do not necessarily represent the official views of the FDA nor FDA’s Office of Orphan Products Development.

## COMPETING INTEREST STATEMENT

DW received advisory board consulting fees from Mitsubishi Tanabe Pharma America, Biogen, and Clene Nanomedicine; KG received funding from the ALS Association, Amylyx, Massachusetts General Hospital, and Zydus; SAD received funding from Biogen, Edgewise Therapeutics, Woolsey Pharmaceuticals, Uniqure Biopharma, Coya Therapeutics, the ALS Foundation, the Peripheral Neuropathy Foundation, and Massachusetts General Hospital and speaking honoraria from Biogen and AANEM; AS received funding from Biogen, Tanabe Pharma America, Amylyx, and the ALS Association; AXO received funding from the Henry Jackson Foundation and Washington University St. Louis; THP received funding from the ALS Association, Amylyx, Mitsubishi Tanabe Pharma America, the State of Pennsylvania, MDA, Massachusetts General Hospital, Duke University, Acellis, and Coya Therapeutics and consulting fees or speaking honoraria from Alpha Insights, Evidera, Decile Ten LLC, MJH Holdings, IQVIA, P Value, Vindico Medical Education, Projects in Knowledge, Impact Education, Kaplan Inc., Leidos, and Neurovigil and advisory board consulting fees from Mitsubishi Tanabe Pharma, Amylyx, Novartis, Biogen, AB Bio, Cytokinetics, and Alexion; GH received speaking honoraria from MTPA, Alexion, and Argenx. All other authors have no relevant disclosures to report.

## Notes

### Author Declarations

The data and biosample collection for this study are approved by the University of Minnesota IRB as the IRB of record. The current IRB number is sIRB 00019204.

