## Supplemental Figures for "Predictive ALS survival using ALSFRS-R slope & NfL: insights from the ALS/MND Natural History Consortium data and biofluid collection"

^1^ Division of Biostatistics & Health Data Science, School of Public Health, University of Minnesota; ^2^ Department of Laboratory Medicine and Pathology, University of Minnesota; ^3^ Masonic Institute for the Developing Brain, University of Minnesota; ^4^ Department of Neurology, University of Minnesota Medical School; ^5^ Department of Neurology, University of Florida; ^6^ Neurology, Temple University; ^7^ Department of Neurology, Saint Louis University; ^8^ Neurology, Lahey Hospital & Medical Center; ^9^ Department of Neurology, University of Pittsburgh; ^10^ Department of Neurology, Northwestern University; ^11^ Neurology, Providence Brain and Spine Institute; ^12^ Neurology, Henry Ford Health; ^13^ Department of Neurology, Virginia Commonwealth University; ^14^ Center for Innovation and Bioinformatics, Massachusetts General Hospital.

*Corresponding author: Andres Arguedas

Supplemental Figures

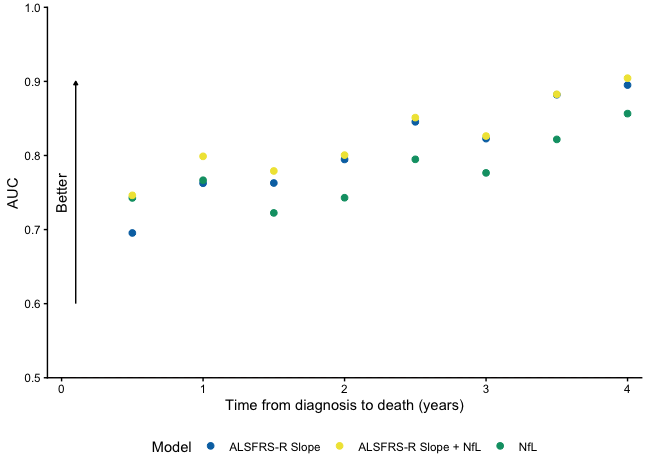

Fig. S1. Dynamic AUC for survival prediction models using baseline NfL values and ALSFRS-R related variables every 6 months after diagnosis (time 0). Models including the baseline NfL value and the ALSFRS-R average rate of change are presented.
